# Recent Intake of Direct Oral Anticoagulants and Acute Ischemic Stroke: Real World Data from a Comprehensive Stroke Center

**DOI:** 10.1101/2025.08.01.25332629

**Authors:** Doreen Pommeranz, Nicole Lehr, Jordi Kühne Escolà, Bastian Brune, Philipp Dammann, Yan Li, Cornelius Deuschl, Michael Forsting, Clemens Kill, Christoph Kleinschnitz, Martin Köhrmann, Benedikt Frank

## Abstract

**Background:** Deciding on intravenous thrombolysis (IVT) in acute ischemic stroke (AIS) patients with reported recent direct oral anticoagulant (DOAC) intake remains challenging due to concerns about hemorrhagic risk and limited real-world evidence. This study characterizes AIS patients with recent DOAC use treated at a comprehensive stroke center, routinely performing quantitative anticoagulation testing.

**Methods:** In this retrospective, registry-based study, we analyzed clinical and procedural data from AIS patients with recent DOAC intake and calibrated anti-factor IIa/Xa activity measured within three hours of admission. Patients were treated at the University Hospital Essen between March 2017 and October 2023.

**Results:** Among 469 included patients, anti-factor IIa/Xa activity was ≤30 ng/ml in 28%, >30–≤50 ng/ml in 9%, >50–≤75 ng/ml in 9%, >75–≤100 ng/ml in 9% and >100 ng/ml in 45%. Lower DOAC levels correlated with severe stroke symptoms at admission (*ρ* = −0.263, *p* <0.001). IVT was administered to 33.5% of patients with DOAC levels ≤50 ng/ml, compared to only 4% among those with levels >50 ng/ml, the majority of whom received prior reversal with idarucizumab. Symptomatic intracranial haemorrhage (sICH) occurred in 4% of IVT-treated and 1% of non-IVT-treated patients, without association to anticoagulation status.

**Conclusion:** A considerable proportion of AIS patients with recent DOAC intake exhibited minimal or no anticoagulant activity at presentation. Those with the lowest levels also showed highest stroke severity. IVT was safe across all DOAC level groups, with low and comparable sICH rates. These findings support the rationale for a randomized trial evaluating IVT without prior DOAC level testing.

## Introduction

While clinical trials have underscored the safety and efficacy of direct oral anticoagulants (DOACs) in stroke prevention, real-world data examining their impact on acute stroke therapy pathways remain limited.

DOACs have shown non-inferiority or superiority to vitamin K antagonists in preventing thromboembolic events, with a more favorable bleeding profile (Poller, Jespersen et al. 2009, Granger, Alexander et al. 2011, Patel, Mahaffey et al. 2011, Giugliano, Ruff et al. 2013). As a result, their use for stroke prevention in patients with non-valvular atrial fibrillation (NVAF) has steadily increased (Steinberg, Gao et al. 2017). Fixed dosing regimes, fewer dieatary and drug interactions and no requirement of drug monitoring are assumed to enhance therapy adherence.

Despite these advantages, 1-2% of patients on DOACs for NVAF experience an acute ischemic stroke (AIS) each year (Seiffge, De Marchis et al. 2020, Vinding, Butt et al. 2022). Among patients eligible for intravenous thrombolysis (IVT), 10–20% are on DOACs (Kupper, Feil et al. 2021, Meinel, Branca et al. 2021, Kam, Holmes et al. 2022). However, IVT is often withheld in these patients due to concerns about an increased risk of hemorrhage (Xian, Liang et al. 2012). According to current European Stroke Organisation (ESO) guidelines, DOAC intake within 48 hours prior to stroke onset constitutes a relative contraindication to IVT (Berge, Whiteley et al. 2021). Consequently, many patients are excluded from thrombolysis based solely on reported intake time, despite considerable inter-and intra-individual variability in DOAC pharmacokinetics and pharmacodynamics (Seiffge, Hooff et al. 2015, Touze, Gruel et al. 2018, Shahjouei 2020). Calibrated anti-factor IIa/Xa activity assays have emerged as reliable indicators of anticoagulant activity (Shahjouei, Tsivgoulis et al. 2020, Berge, Whiteley et al. 2021). Nevertheless, current safety thresholds for IVT eligibility are primarily based on expert opinion and limited pilot data (Seiffge, Meinel et al. 2021). According to the summaries of product characteristics (SmPC) for alteplase and tenecteplase, IVT may be considered if coagulation tests are below the respective upper limit of normal (EMA 2024). However, the use of such tests is constrained by feasibility, costs and accessibility, limiting their routine implementation to specific conditions and selected, experienced centers (Luca, Oliva et al. 2023, Kristoffersen, Seiffge et al. 2024).

This study provides a detailed clinical profile of AIS patients with reported DOAC use, including routine measurement of calibrated anti-factor IIa/Xa activity at the time of hospital admission.

## Methods

### Study design

This retrospective, single-center, observational cohort study was conducted at University Hospital Essen. The study protocol was reviewed and approved by the local ethics committee (18-8408-BO). Due to the retrospective nature of the study, formal patient consent was not required. Nonetheless, all data were handled in accordance with institutional ethical standards, national data protection regulations and the Declaration of Helsinki.

### Patient population and data acquisition

We included data from AIS patients with reported recent intake of apixaban, edoxaban, rivaroxaban or dabigatran who presented to the emergency department between March 2017 and October 2023. Patients were eligible if they met the following criteria:

1. Clinical and/or imaging-based diagnosis of AIS (including transient ischemic attack); computed tomography (CT) was the preferred modality.
2. Documented current DOAC therapy and reported recent intake.
3. Measurement of calibrated anti-factor IIa (dabigatran) or anti-factor Xa (apixaban, edoxaban, rivaroxaban) activity within 3 hours of hospital admission.
4. Age ≥18 years.

Exclusion criteria were:

1. Time from symptom onset to admission >72 hours.
2. Stroke mimic, intracranial hemorrhage (ICH) or any alternative primary diagnosis.

Data were sourced from the institutional stroke registry. Laboratory results were exported from the local information system and merged with clinical data. Structured clinical information including the National Institutes of Health Stroke Scale (NIHSS) and the modified Rankin Scale (mRS) was entered by treating neurologists. Follow-up neuroimaging was performed within 24 hours after IVT and/or endovascular thrombectomy (ET) or earlier in the event of new or worsening neurological deficits. All imaging was independently reviewed by a neurologist and a neuroradiologist. ICH was classified according to the Heidelberg Bleeding Classification (von Kummer, Broderick et al. 2015). Symptomatic ICH (sICH) was defined based on ECASS III criteria as a hemorrhage causing neurological deterioration (≥4-point increase in NIHSS) or resulting in death (Hacke, Kaste et al. 2008).

### Laboratory Testing

At the University Hospital Essen, calibrated anti-factor IIa/Xa activity is routinely measured in all AIS patients reported to be on DOACs. Blood samples for coagulation testing are drawn immediately upon admission, typically in parallel with NIHSS assessment or neuroimaging.

According to institutional protocols (Figure 1), IVT with 0.9 mg/kg alteplase is administered to patients with DOAC plasma levels <50 ng/ml, irrespective of the time of last DOAC intake. For patients on direct factor Xa inhibitors with levels between 50-100 ng/ml, IVT is considered based on individualized risk-benefit assessment, with ET preferred as the first-line treatment where appropriate.

**Figure 1.**
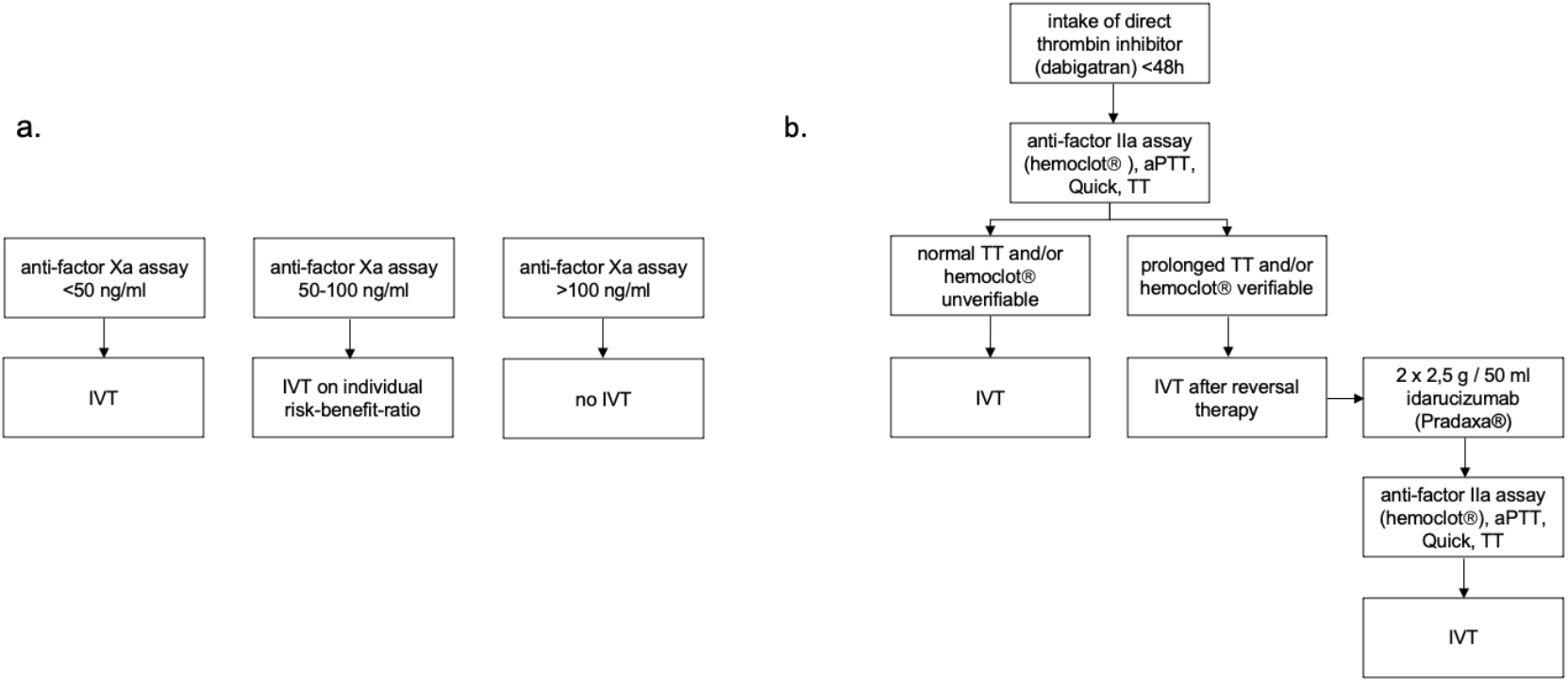
Institutional guidelines for the acute stroke management of patients under therapy with (a.) direct factor Xa inhibitors and (b.) direct thrombin inhibitors. Abbreviations: aPTT = activated partial thromboplastin time; IVT = intravenous thrombolysis; TT = thrombin time.

Patients on the direct thrombin inhibitor dabigatran represent a special subgroup due to the availability of the specific reversal agent idarucizumab. To guide reversal therapy decisions, calibrated anti-factor IIa activity (HEMOCLOT®), activated partial thromboplastin time (aPTT) and thrombin time (TT) are measured. A normal TT -the most sensitive global test - excludes clinically relevant dabigatran activity (Levy, Ageno et al. 2016). In cases of normal TT or undetectable HEMOCLOT®, IVT is administered without reversal. If TT is prolonged or HEMOCLOT® is quantifiable, idarucizumab 2 × 2.5 g is administered, followed by reassessment of HEMOCLOT®, aPTT and TT. IVT is then initiated without additional delay for laboratory confirmation.

### Statistical analysis

All statistical analyses were performed using SPSS Statistics, version 29 (IBM Corp., Armonk, NY, USA). Figures were generated using Microsoft Excel, version 16.89.1 (Microsoft Corp., Redmond, WA, USA). The cohort was stratified into five groups based on commonly referenced DOAC plasma level thresholds. The widely accepted threshold for “safe treatment” with IVT or surgery is <30 ng/ml (Ahmed, Audebert et al. 2019). The French Society of Vascular Neurology recommends consideration of IVT at DOAC plasma levels <50 ng/ml (Touze, Gruel et al. 2018), a threshold supported by a German cohort study reporting only one sICH among 261 patients (Marsch, Macha et al. 2019). Prior studies and institutional guidelines often define a DOAC plasma level of 100 ng/ml as the upper treshold for IVT eligibility. Two single-center studies from Basel (Switzerland) and Erlangen (Germany) investigated 18 and 24 AIS patients, respectively, who received IVT despite DOAC plasma levels up to 100 ng/ml. No sICH was observed in the Basel cohort and only one case was reported in the Erlangen cohort (Seiffge, Traenka et al. 2017, Marsch, Macha et al. 2019).

Descriptive statistics were used to summarize demographics, treatment and outcomes. Continuous variables are presented as medians with interquartile ranges (IQR), categorical variables as absolute and relative frequencies. For subgroup analyses, the cohort was dichotomized into patients without anticoagulant activity (anti-factor IIa/Xa activity ≤30 ng/ml) and those with anticoagulant activity (anti-factor IIa/Xa activity >30 ng/ml). Fisher’s exact test and the Mann-Whitney U test were used to compare categorical and continuous variables, respectively. Statistical significance was set at p <0.05 (two-tailed). Additional subgroup analyses compared IVT- and non-IVT-treated patients across DOAC levels. Spearman’s rank correlation was used to assess the association between DOAC level and stroke severity at admission.

## Results

Between March 2017 and October 2023 a total of 742 patients with reported recent DOAC intake were admitted to the University Hospital Essen due to suspected acute stroke (Figure 2). Among these, 187 (25%) were diagnosed with stroke mimics, most commonly seizures (26%) and infections (22%). A clinical diagnosis of stroke was confirmed in 555 patients (75%), with neuroimaging identifying intracranial hemorrhage (ICH) in 86 cases (15%), including 22 traumatic and 64 non-traumatic hemorrhages. Ultimately, 469 of 555 patients (85%) were diagnosed with acute ischemia.

**Figure 2.**
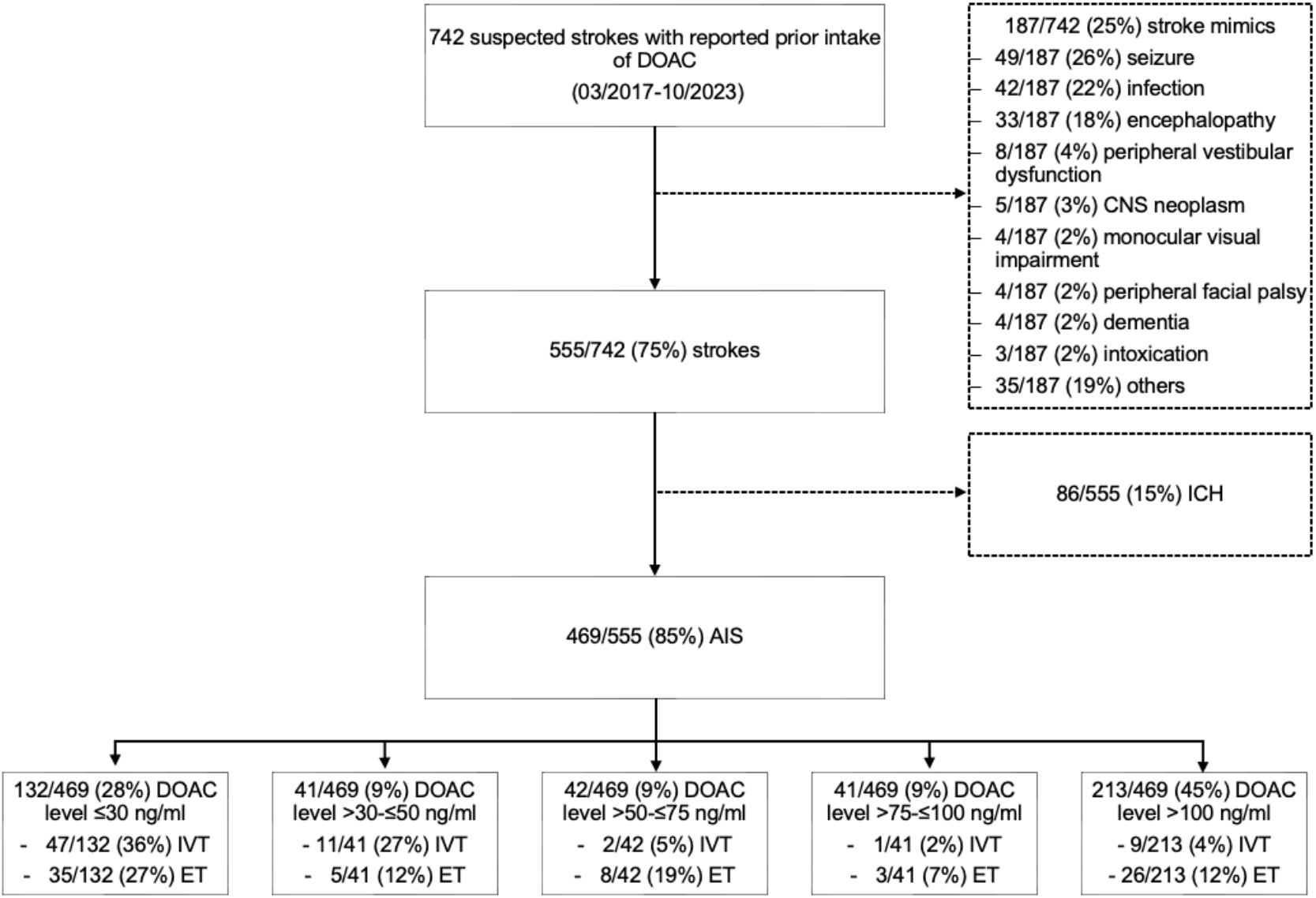
Flowchart of study population inclusion process. Abbreviations: AIS = acute ischemic stroke; CNS = central nervous system; ET = endovascular therapy; ICH = intracranial hemorrhage; IVT = intravenous thrombolysis.

Among these 469 AIS patients, DOAC plasma level was ≤30 ng/ml in 132 (28%) patients, >30-≤50 ng/ml in 41 (9%), >50-≤75 ng/ml in 42 (9%), >75-≤100 ng/ml in 41 (9%), and >100 ng/ml in 213 (45%).

### Baseline characteristics

The median age of the cohort was 81 years (IQR 74–86) and 54% were female (Table 1). Comorbidities were common and comparably distributed across DOAC level subgroups. Prior stroke was reported in 50% of patients with DOAC levels >100 ng/ml (106/213). Renal impairment (serum creatinine ≥1.2 mg/dl) was present in 36% of all patients, with no clear association to DOAC level.

**Table 1.** Baseline characteristics, procedural data and outcomes stratified by DOAC plasma level.

|  | Overall<br>(n=469) | DOAC<br>level ≤30<br>ng/ml<br>(n=132,<br>28%) | DOAC<br>level >30-<br>≤50 ng/ml<br>(n=41, 9%) | DOAC<br>level >50-<br>≤75 ng/ml<br>(n=42, 9%) | DOAC<br>level >75-<br>≤100 ng/ml<br>(n=41, 9%) | DOAC<br>level >100<br>ng/ml<br>(n=213,<br>45%) |
| --- | --- | --- | --- | --- | --- | --- |
| **Demographics and functional status** |  |  |  |  |  |  |
| Age (in years) | 81 (74-86) | 81 (71-86) | 80 (70-86) | 82 (78-86) | 84 (71-88) | 81 (75-86) |
| Females | 254 (54%) | 77 (58%) | 21 (51%) | 21 (50%) | 21 (51%) | 114 (54%) |
| NIHSS at admission | 6 (2-13) | 10 (5-18) | 6 (1-11) | 7 (3-14) | 5 (2-12) | 4 (2-11) |
| mRS ≤2 prior to admission | 276 (59%) | 81 (61%) | 27 (66%) | 26 (62%) | 24 (59%) | 118 (55%) |
| **Comorbidities** |  |  |  |  |  |  |
| Arterial hypertension | 345 (74%) | 99 (75%) | 30 (73%) | 35 (83%) | 30 (73%) | 151 (71%) |
| Diabetes | 135 (29%) | 32 (24%) | 10 (24%) | 13 (31%) | 12 (29%) | 68 (32%) |
| Prior Stroke | 206 (44%) | 50 (38%) | 10 (24%) | 22 (52%) | 18 (44%) | 106 (50%) |
| Renal insufficiency* | 167 (36%) | 45 (34%) | 11 (27%) | 10 (24%) | 17 (42%) | 84 (39%) |
| **Indication for DOAC** |  |  |  |  |  |  |
| NVAF | 423 (90%) | 114 (86%) | 33 (81%) | 41 (98%) | 40 (98%) | 195 (92%) |
| Deep vein thrombosis | 13 (3%) | 4 (3%) | 1 (2%) | 1 (2%) | 1 (2%) | 6 (3%) |
| Pulmonary embolism | 15 (3%) | 4 (3%) | 6 (15%) | - | - | 5 (2%) |
| Peripheral artery disease | 4 (0.9%) | 3 (2%) | - | - | - | 1 (0.5%) |
| Coagulopathy | 12 (3%) | 6 (5%) | 1 (2%) | - | - | 5 (2%) |
| LAAO/Valvular transplant | 2 (0.4%) | 1 (0.8%) | - | - | - | 1 (0.5%) |
| **Type of DOAC** |  |  |  |  |  |  |
| Apixaban | 248 (53%) | 58 (44%) | 21 (51%) | 22 (52%) | 22 (54%) | 125 (59%) |
| Dabigatran | 39 (8%) | 7 (5%) | 5 (12%) | 5 (12%) | 4 (10%) | 18 (9%) |
| Edoxaban | 99 (21%) | 39 (30%) | 8 (20%) | 7 (17%) | 12 (29%) | 33 (16%) |
| Rivaroxaban | 83 (18%) | 28 (21%) | 7 (17%) | 8 (19%) | 3 (7%) | 37 (17%) |
| **Blood sample analysis** |  |  |  |  |  |  |
| INR | 1.09 (1.03-1.19) | 1.04 (0.99-1.10) | 1.04 (1.00-1.11) | 1.09 (1.04-1.17) | 1.10 (1.03-1.18) | 1.15 (1.06-1.32) |
| aPTT | 26.10 (23.80-29.15) | 23.70 (22.20-25.60) | 26.00 (23.90-28.00) | 25.6 (24.00-28.90) | 26.30 (24.10-28.70) | 28.00 (25.65-31.65) |
| **Procedural data** |  |  |  |  |  |  |
| DTN (in minutes)** | 53 (40-72) | 50 (40-73) | 54 (47-72) | 65 (-) | 57 (-) | 52 (33-96) |
| DTGP (in minutes) | 67 (46-86) | 75 (55-97) | 30 (21-43) | 78 (40-114) | 65 (-) | 66 (51-80) |
| Length of hospital stay (in days) | 4 (2-6) | 6 (4-10) | 4 (3-5) | 5 (3-7) | 4 (1-5) | 4 (3-8) |
| **Therapy** |  |  |  |  |  |  |
| IVT | 70 (15%) | 47 (36%) | 11 (27%) | 2 (5%) | 1 (2%) | 9 (4%) |
| ET | 77 (16%) | 35 (27%) | 5 (12%) | 8 (19%) | 3 (7%) | 26 (12%) |
| PPSB | 2 (0.4%) | 1 (0.8%) | - | 1 (2%) | - | - |
| Idarucizumab | 9 (2%) | 2 (2%) | 2 (5%) | - | - | 5 (2%) |

| **Outcomes** |  |  |  |  |  |  |
| --- | --- | --- | --- | --- | --- | --- |
| NIHSS at discharge | 3 (1-9) | 5 (2-20) | 3 (1-9) | 4 (2-13) | 3 (1-7) | 3 (1-7) |
| ΔNIHSS*** | 1 (0-4) | 1 (-2-5) | 0 (-1-4) | 1 (0-4) | 1 (0-5) | 1 (0-3) |
| mRS >2 at discharge | 281 (60%) | 92 (70%) | 21 (51%) | 28 (67%) | 21 (51%) | 120 (56%) |
| ICH | 18 (4%) | 5 (4%) | 2 (5%) | 2 (5%) | 2 (5%) | 7 (3%) |
| - Initial imaging | 3 (0.6%) | 1 (0.8%) | - | - | - | 2 (0.9%) |
| - Follow-up imaging**** | 15 (3%) | 4 (3%) | 2 (5%) | 2 (5%) | 2 (5%) | 5 (2%) |
| sICH | 8 (2%) | 2 (2%) | 2 (5%) | 1 (2%) | - | 3 (1%) |
| In-hospital mortality | 59 (13%) | 27 (20%) | 6 (15%) | 7 (17%) | 2 (5%) | 17 (8%) |

Data are n (%) or median (IQR).
\*renal insufficiency = serum creatinine ≥ 1.2 mg/dl
\*\*one missing due to externally performed IVT
\*\*\* $\Delta$ NIHSS = NIHSS at admission - NIHSS at discharge
\*\*\*\*first sight of ICH

The predominant indication for DOAC therapy was NVAF in 90% of cases, followed by pulmonary embolism (3%) and deep vein thrombosis (3%). Apixaban was the most frequently used agent (53%), followed by edoxaban (21%), rivaroxaban (18%) and dabigatran (8%).

Stroke severity at admission, measured by the NIHSS, had a median value of 6. A significant inverse correlation was observed between DOAC plasma levels and NIHSS scores (Spearman’s ρ = −0.263, p <0.001), indicating that lower anticoagulation levels were associated with more severe strokes (Figure 3).

**Figure 3.**
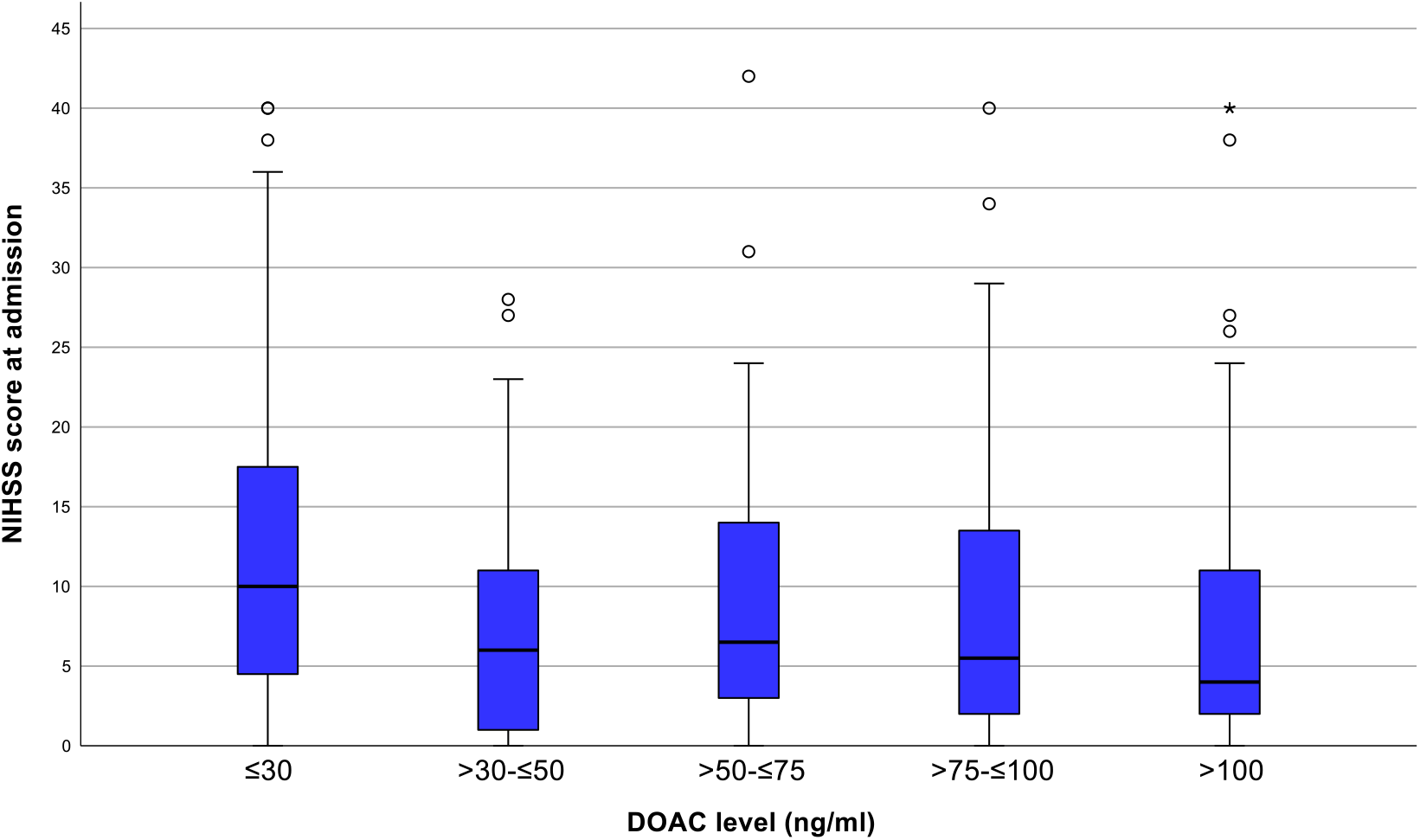
Stroke severity at admission across DOAC plasma level groups. Boxplots displaying NIHSS scores at admission stratified by DOAC plasma level groups (≤30 ng/ml, >30–≤50 ng/ml, >50–≤75 ng/ml, >75–≤100 ng/ml, and >100 ng/ml). Boxes represent interquartile ranges (IQR), horizontal lines indicate medians, whiskers denote 1.5 x IQR and circles represent outliers.

### Procedural data

IVT was administered in 70 of 469 patients (15%), with 24 of these (34%) also undergoing endovascular thrombectomy (ET). The ET rate ranged from 7% to 27% across DOAC level groups, while IVT administration showed a near-linear decline with increasing DOAC levels (Tables 1 and 2).

**Table 2.**
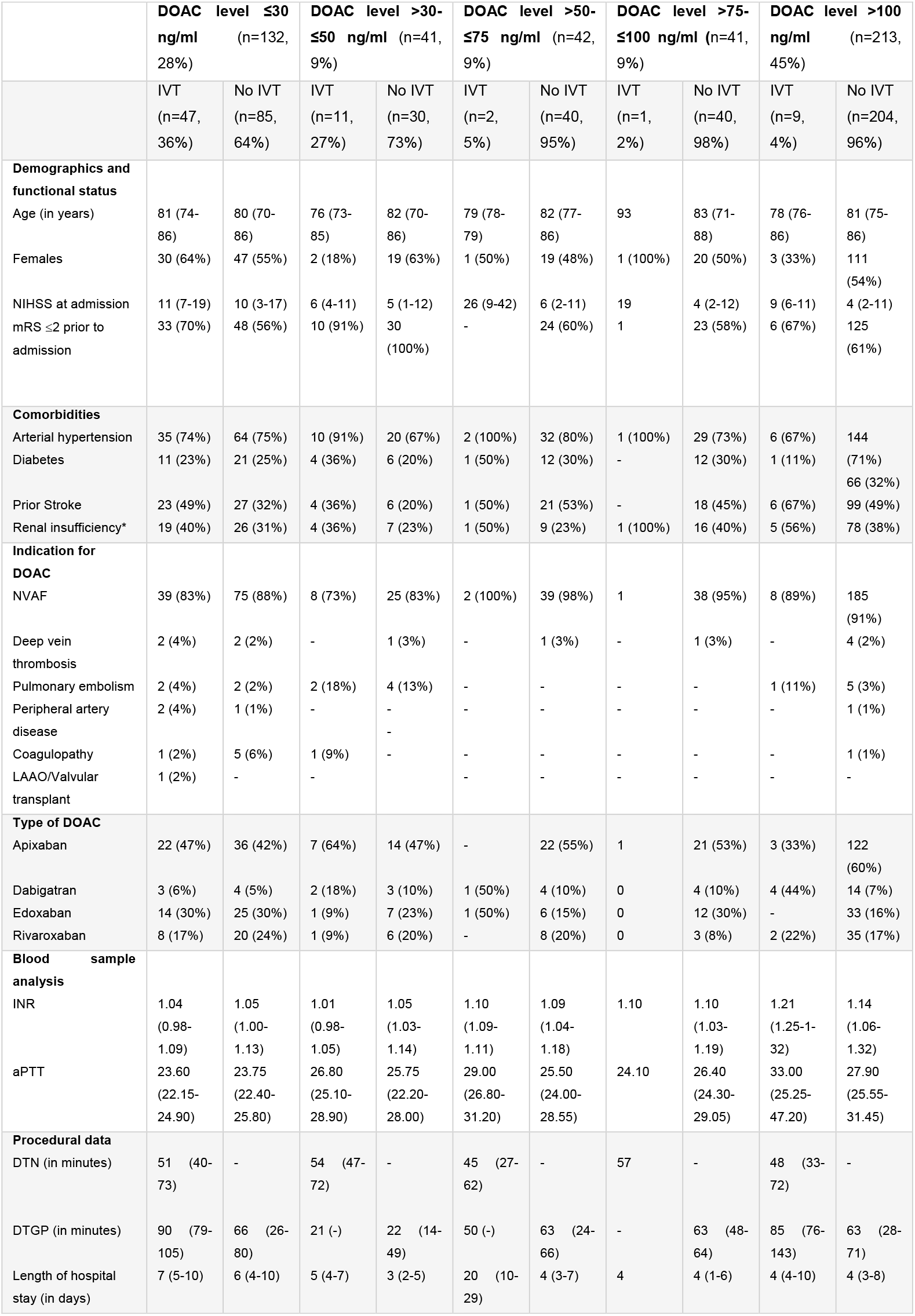
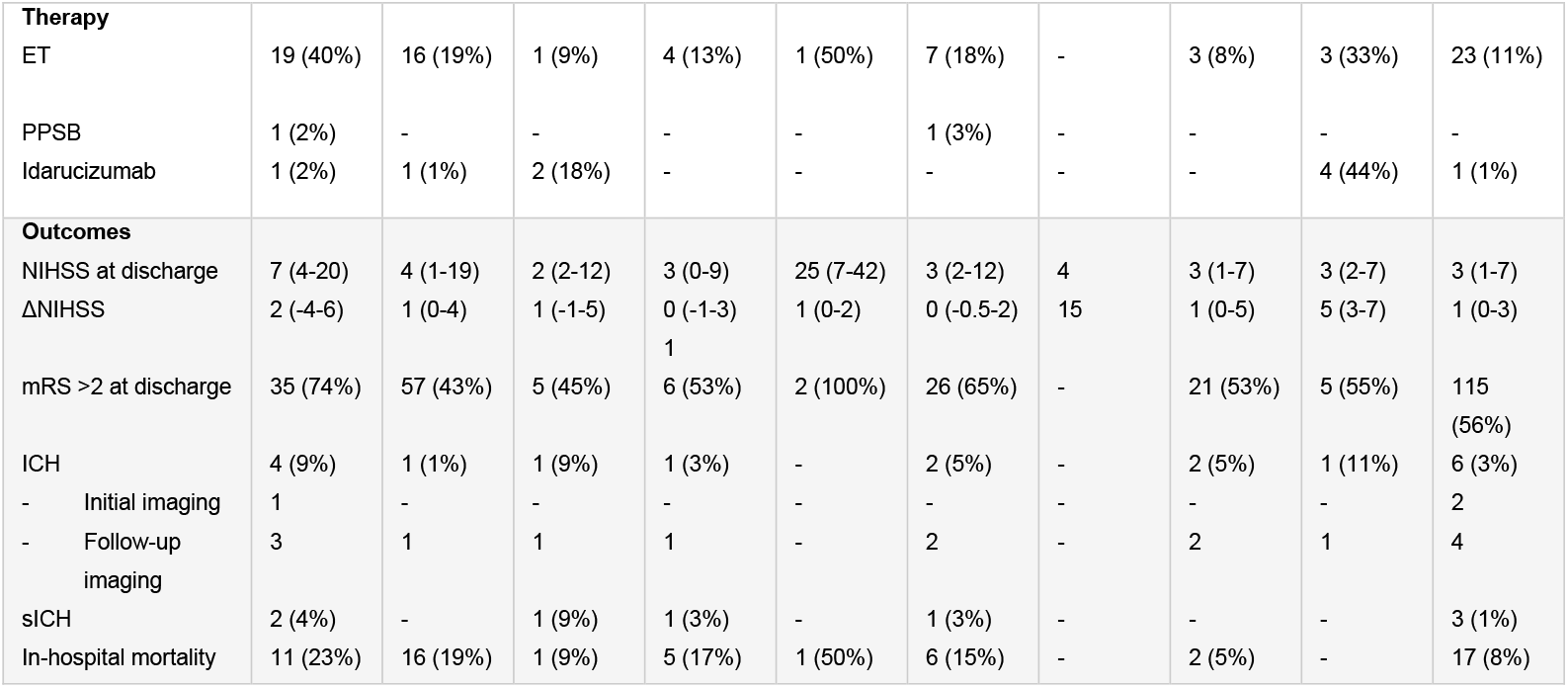
IVT-treated patients compared to non-IVT-treated patients, stratified by DOAC plasma level.

According to institutional protocols consistent with European guidelines, 173 patients (37%) had DOAC levels <50 ng/ml, considered safe for IVT. Additionally, 83 patients (18%) fell within the “gray zone” (50–100 ng/ml), where IVT is considered based on individualized risk assessment. Despite these findings, only 70 patients (15%) received IVT, predominantly those with DOAC levels ≤30 ng/ml (n = 47). IVT was rarely used in patients with levels >30 ng/ml (n = 23) and often preceded by reversal therapy. The median door-to-needle (DTN) time in the cohort was 53 minutes (IQR 40–72).

### Safety Outcomes

ICH was observed in 18 of 469 patients (4%). The most frequent subtype was subarachnoid hemorrhage (SAH, n = 7), followed by hemorrhagic infarction type 1 (HI1, n = 4), hemorrhagic infarction type 2 (HI2, n = 2), parenchymal hematoma type 2 (PH2, n = 2), and intraventricular hemorrhage (IVH, n = 1). Three cases were detected on initial imaging, while the remaining 15 were identified during follow-up.

Symptomatic ICH (sICH) occurred in 8 patients (2%), with no evident association to DOAC level or IVT administration (sICH rate: 3/70 in the IVT group, 5/399 in the non-IVT group). Among 17 patients with DOAC levels >30 ng/ml who received IVT without reversal, no cases of sICH occurred in those with levels >50 ng/ml (Table 2). The most frequent type of sICH was SAH (n = 3), classified according to the Heidelberg Bleeding Classification.

### Anticoagulation status

To investigate differences between patients without anticoagulant activity and those with anticoagulant activity (defined as a calibrated anti-factor IIa/Xa activity ≤30 ng/ml respectively >30 ng/ml), a dichotomization of the cohort was performed (Table 3). Demographics, comorbidities, DOAC indication and agent and procedural parameters were similar between groups (all p ≥0.05).

**Table 3.**
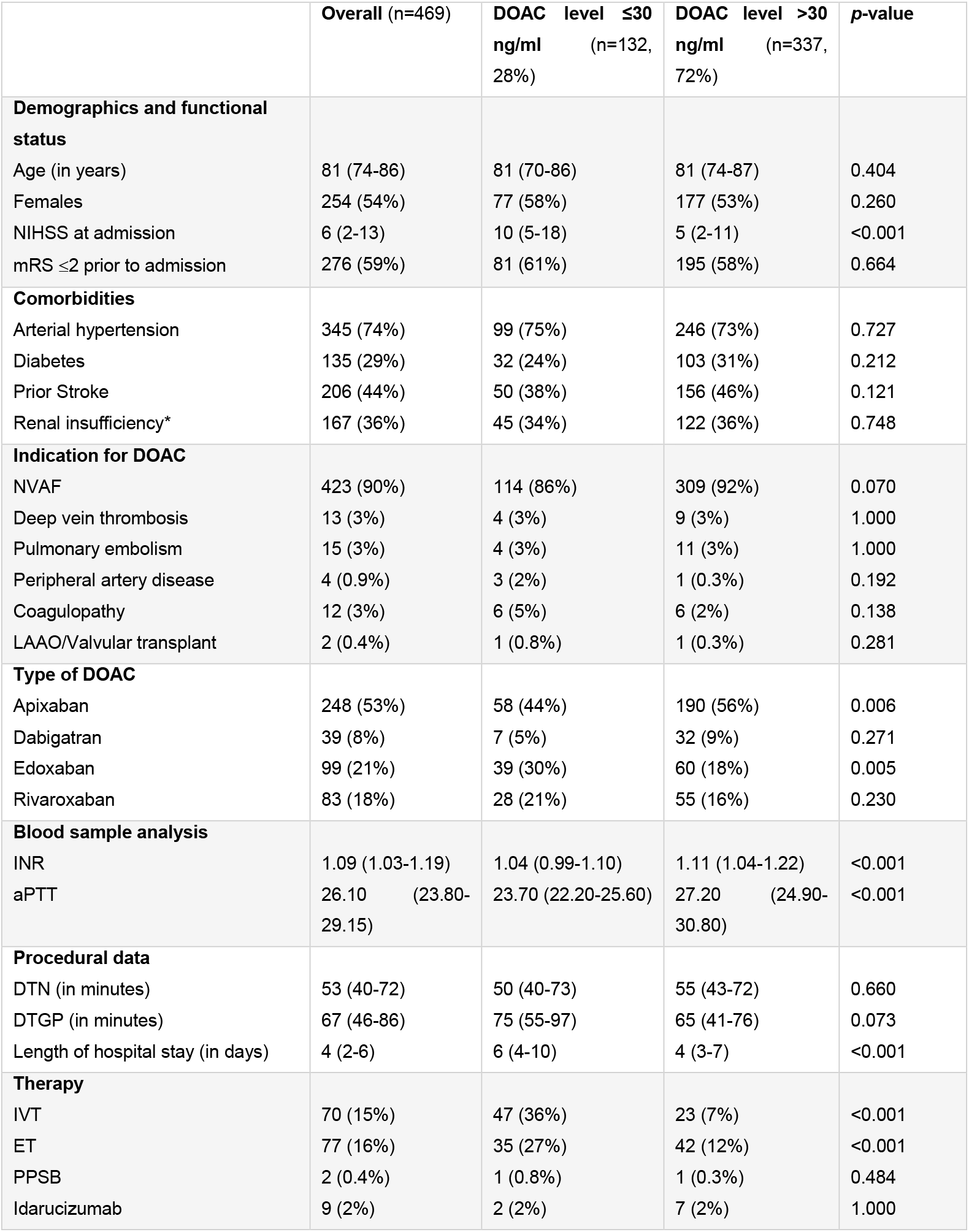
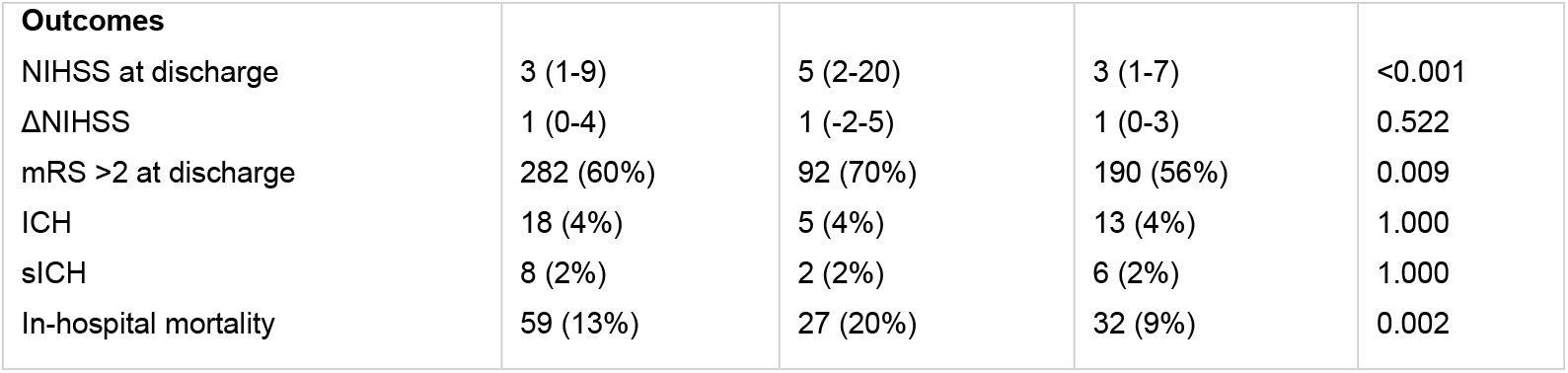
Patients without anticoagulant activity (Calibrated anti-factor IIa/Xa activity ≤30 ng/ml) compared to patients with anticoagulant activity (Calibrated anti-factor IIa/Xa activity >30 ng/ml).

| | Overall (n=469) | DOAC level $\leq 30$<br>ng/ml (n=132,<br>28%) | DOAC level $>30$<br>ng/ml (n=337,<br>72%) | p-value |
| --- | --- | --- | --- | --- |
| **Demographics and functional status** |  |  |  |  |
| Age (in years) | 81 (74-86) | 81 (70-86) | 81 (74-87) | 0.404 |
| Females | 254 (54%) | 77 (58%) | 177 (53%) | 0.260 |
| NIHSS at admission | 6 (2-13) | 10 (5-18) | 5 (2-11) | <0.001 |
| mRS $\leq 2$ prior to admission | 276 (59%) | 81 (61%) | 195 (58%) | 0.664 |
| **Comorbidities** |  |  |  |  |
| Arterial hypertension | 345 (74%) | 99 (75%) | 246 (73%) | 0.727 |
| Diabetes | 135 (29%) | 32 (24%) | 103 (31%) | 0.212 |
| Prior Stroke | 206 (44%) | 50 (38%) | 156 (46%) | 0.121 |
| Renal insufficiency* | 167 (36%) | 45 (34%) | 122 (36%) | 0.748 |
| **Indication for DOAC** |  |  |  |  |
| NVAF | 423 (90%) | 114 (86%) | 309 (92%) | 0.070 |
| Deep vein thrombosis | 13 (3%) | 4 (3%) | 9 (3%) | 1.000 |
| Pulmonary embolism | 15 (3%) | 4 (3%) | 11 (3%) | 1.000 |
| Peripheral artery disease | 4 (0.9%) | 3 (2%) | 1 (0.3%) | 0.192 |
| Coagulopathy | 12 (3%) | 6 (5%) | 6 (2%) | 0.138 |
| LAAO/Valvular transplant | 2 (0.4%) | 1 (0.8%) | 1 (0.3%) | 0.281 |
| **Type of DOAC** |  |  |  |  |
| Apixaban | 248 (53%) | 58 (44%) | 190 (56%) | 0.006 |
| Dabigatran | 39 (8%) | 7 (5%) | 32 (9%) | 0.271 |
| Edoxaban | 99 (21%) | 39 (30%) | 60 (18%) | 0.005 |
| Rivaroxaban | 83 (18%) | 28 (21%) | 55 (16%) | 0.230 |
| **Blood sample analysis** |  |  |  |  |
| INR | 1.09 (1.03-1.19) | 1.04 (0.99-1.10) | 1.11 (1.04-1.22) | <0.001 |
| aPTT | 26.10 (23.80-29.15) | 23.70 (22.20-25.60) | 27.20 (24.90-30.80) | <0.001 |
| **Procedural data** |  |  |  |  |
| DTN (in minutes) | 53 (40-72) | 50 (40-73) | 55 (43-72) | 0.660 |
| DTGP (in minutes) | 67 (46-86) | 75 (55-97) | 65 (41-76) | 0.073 |
| Length of hospital stay (in days) | 4 (2-6) | 6 (4-10) | 4 (3-7) | <0.001 |
| **Therapy** |  |  |  |  |
| IVT | 70 (15%) | 47 (36%) | 23 (7%) | <0.001 |
| ET | 77 (16%) | 35 (27%) | 42 (12%) | <0.001 |
| PPSB | 2 (0.4%) | 1 (0.8%) | 1 (0.3%) | 0.484 |
| Idarucizumab | 9 (2%) | 2 (2%) | 7 (2%) | 1.000 |

| **Outcomes** |  |  |  |  |
| --- | --- | --- | --- | --- |
| NIHSS at discharge | 3 (1-9) | 5 (2-20) | 3 (1-7) | <0.001 |
| ΔNIHSS | 1 (0-4) | 1 (-2-5) | 1 (0-3) | 0.522 |
| mRS >2 at discharge | 282 (60%) | 92 (70%) | 190 (56%) | 0.009 |
| ICH | 18 (4%) | 5 (4%) | 13 (4%) | 1.000 |
| sICH | 8 (2%) | 2 (2%) | 6 (2%) | 1.000 |
| In-hospital mortality | 59 (13%) | 27 (20%) | 32 (9%) | 0.002 |

Patients without anticoagulant activity presented with significantly severe stroke symptoms at admission (*p* <0.001) and were more likely to experience longer hospital stays (*p* <0.001), higher in-hospital mortality (*p* <0.001) and worse functional outcome (*p* = 0.009). In contrast, no significant difference was observed in the prevalence of sICH (*p* = 1.000). Although global coagulation parameters are not specific for DOAC activity, both INR and aPTT were significantly higher in patients with higher DOAC levels (p <0.001).

## Discussion

The key findings of this study are: (1) a substantial proportion of AIS patients with reported recent DOAC intake indicated no anticoagulant activity at the time of admission; (2) IVT was rarely performed despite laboratory eligibility; and (3) low DOAC plasma levels were associated with more severe strokes, yet sICH rates remained low across all subgroups.

Approximately one-third of patients with reported recent DOAC intake showed no anticoagulant activity at admission. This suggests a significant gap between prescribed therapy and effective anticoagulant levels. Despite the routine availability of quantitative DOAC testing at our center, treatment decisions frequently reflected a conservative approach. This hesitation may be rooted in the historical exclusion of DOAC-treated patients from pivotal IVT trials. However, emerging evidence increasingly supports the safety of IVT in selected patients on DOACs, particularly when decisions are informed by calibrated anti-factor IIa/Xa assays (Kam, Holmes et al. 2022, Matusevicius, Saflund et al. 2025).

Notably, no increased risk of sICH was observed in patients with DOAC levels >30 ng/ml. Rates of sICH were comparable between patients without (2%) and patients with anticoagulant activity (2%) and between IVT-treated (4%) and non-IVT-treated (1%) groups. Among 17 patients indicating DOAC levels >30 ng/ml who underwent IVT without reversal, only one developed sICH. These findings are consistent with prior observational studies reporting low hemorrhagic risk in this population (Ghannam, AlMajali et al. 2023). Furthermore, patients with low DOAC levels had significantly higher stroke severity, increased in-hospital mortality and poorer functional outcomes, aligning with previous reports that adequate DOAC therapy mitigates stroke severity without significantly elevating bleeding risk (Meinel, Branca et al. 2021, Rizos, Meid et al. 2022, Meinel, Wilson et al. 2023).

The median door-to-needle (DTN) time of 53 minutes in our cohort represents a substantial delay compared to our institutional benchmark of 30 minutes (Chae, Vössing et al. 2023). This prolongation was primarily attributable to the turnaround time of ∼30 minutes for DOAC plasma level testing. The longest DTN times were observed in patients with DOAC levels 50-75 ng/ml - a range associated with therapeutic ambiguity and heightened medicolegal caution. Among DOAC subtypes, the most pronounced delays occurred in dabigatran-treated patients, likely reflecting lower familiarity with idarucizumab use in acute settings. While quantitative testing improves safety, it may delay urgent interventions. Standardized workflows and increased familiarity with reversal protocols could mitigate these delays.

### Limitations and future dircetions

This study has limitations inherent to retrospective analyses. As a single-center observational study, causal inferences cannot be drawn. DOAC levels were measured routinely only at admission, precluding an assessment of dynamic clearance rates. Data on the exact timing and dosing of last DOAC intake were unavailable, limiting interpretation of low plasma levels. Additionally, information on anticoagulation status at prior strokes or changes in DOAC agent and dosage was not systematically recorded. Alternative laboratory methods, such as urine dipstick tests, modified thromboelastography with anti-Xa assays and ecarin clotting time-based cartridges, offer the potential for a more rapid and widely accessible assessment of the anticoagulation status (Seiffge, Meinel et al. 2021).

## Conclusion

A substantial proportion of AIS patients with reported recent DOAC use showed no anticoagulant acitivity at hospital admission. Nevertheless, IVT was underutilized reflecting persistent uncertainty in clinical practice. Importantly, use of IVT was not associated with increased sICH risk in patients with DOAC levels >30 ng/ml. Our findings support the rationale for a randomized controlled trial evaluating IVT in AIS patients with reported DOAC use and without prior DOAC level testing.

## Data Availability

The authors declare that all supporting data are available within the manuscript.

## Acknowledgments

None.

